# Impact of 26 Skin Diseases on the Risk of Non-Small Cell Lung Cancer: A Mendelian Randomization Study Using FinnGen R11 Data

**DOI:** 10.1101/2024.09.05.24313092

**Authors:** Xingyuan Li, Hui Li

## Abstract

**Purpose:** To determine whether genetic predisposition to various skin diseases influences the risk of non-small cell lung cancer (NSCLC) through Mendelian randomization (MR).

**Methods:** Single nucleotide polymorphisms (SNPs) associated with 26 skin diseases were extracted from the FinnGen R11 dataset and underwent rigorous quality control. To evaluate the association between these skin diseases and the risk of non-small cell lung cancer (NSCLC), we applied several analytical methods, including inverse-variance weighted (IVW), MR-Egger regression, weighted median, Simple mode, and Weighted mode. The robustness of the findings was further supported by assessing SNP heterogeneity with the Cochran Q test and evaluating horizontal pleiotropy using the MR-Egger intercept test.

**Results:** Our study revealed that genetically predicted dermatitis herpetiformis (DH) was significantly associated with an elevated risk of squamous cell carcinoma of the lung (SCC). Acne was nominally linked to an increased risk of SCC. Additionally, rhinophyma (RHN), hidradenitis suppurativa (HS), and DH were nominally associated with a higher risk of adenocarcinoma of the lung (ADC). Of the remaining 22 skin diseases analyzed, 7 lacked sufficient instrumental variables to meet inclusion criteria. The other 15 skin diseases showed no statistically significant association with NSCLC.

**Conclusion:** This study ultimately analyzed the relationship between 19 skin diseases and NSCLC at the genetic level, while 7 other skin diseases could not be analyzed due to insufficient instrumental variables. Dermatitis herpetiformis and acne were associated with an increased risk of squamous cell carcinoma of the lung. Additionally, rhinophyma, hidradenitis suppurativa, and dermatitis herpetiformis were associated with an increased risk of adenocarcinoma of the lung.

## 1. Introduction

Lung cancer is one of the most prevalent and deadly cancers worldwide ^[1]^. Non-small cell lung cancer (NSCLC) accounts for about 85% of lung cancer cases, highlighting the need for targeted research and therapies. The most common NSCLC subtypes are adenocarcinoma (ADC) and squamous cell carcinoma (SCC), representing 40% and 25- 30% of cases, respectively ^[2]^. The development and progression of NSCLC are influenced by genetic susceptibility, environmental factors, chronic inflammation, and immune system dysregulation ^[3]^. Despite extensive research, the etiology and pathogenesis of lung cancer remain incompletely understood ^[4]^.

Skin diseases are a group of conditions that affect the skin, hair, nails, and associated tissues. Several studies have already explored the physiological and pathological connections between the skin and lungs, particularly focusing on the shared roles of certain inflammatory pathways in lung and skin inflammation, as well as the relationship between inflammation and lung tumors ^[5]-[7]^, However, there are still many deficiencies in the research on the correlation between specific skin diseases and lung diseases. Previous Mendelian Randomization (MR) studies have also confirmed the association between psoriasis and the risk of lung cancer ^[8]-[9]^, Additionally, a Mendelian randomization study demonstrated a relationship between psoriasis and pulmonary fibrosis ^[10]^. Building on these studies, it is logical to further investigate the relationship between lung cancer and a broader range of skin diseases.

Based on current clinical practice, targeted therapy, immunotherapy, and radiotherapy for lung cancer often lead to varying degrees of adverse reactions in the skin and mucous membranes ^[10]-[11]^. Therefore, in a clinical setting, it is challenging to accurately observe the relationship between skin diseases and lung cancer without these confounding factors. The MR method uses genetic variants as instrumental variables to mitigate confounding factors and provide stronger evidence of causal relationships ^[12]^. Unlike clinical trials, MR can eliminate the influence of social environmental factors and treatment-related effects on study results ^[13]-[14]^, making it an effective tool for studying the genetic links between skin diseases and lung cancer.

## 2. Information and methodology

### 2.1. Sources of information

To utilize the most recent research data and ensure the uniformity of the study population, we chose to obtain the instrumental variables from the June 2024 R11 data of the FinnGen database (index (finngen.fi)). The final selected dataset is detailed in Table1.

### 2.2. Assumptions of the MR study

The Mendelian Randomization (MR) study is grounded in three key assumptions that ensure valid causal inferences ^[15]^. First, the relevance assumption requires that genetic instrumental variables (GIVs) are strongly associated with the exposure factors. Second, the independence assumption requires that GIVs are isolated from any confounders that might impact the correlation between GIVs and outcomes. Third, the exclusion restriction assumption mandates that GIVs influence outcomes solely through the exposure factors, with no direct impact on the outcomes themselves (Fig1). In this study, skin diseases are considered the primary exposure factors, with squamous cell carcinoma (SCC) and adenocarcinoma (ADC) of the lung as the outcomes of interest. The objective is to analyze the potential causal impact of skin diseases on these two major subtypes of non-small cell lung cancer (NSCLC).

### 2.3. Screening for genetic instrumental variables

A stringent threshold of P < 5 × 10⁻⁸ was used to identify single nucleotide polymorphisms (SNPs) significantly associated with exposure factors, qualifying them as genetic instrumental variables (GIVs). This threshold selects only the most statistically significant SNPs, enhancing the reliability of the Mendelian Randomization (MR) analysis. To minimize confounding from linkage disequilibrium (LD), an r² value of 0.001 and a genetic window of 10,000 kilobases (kb) were applied.

The strength of the GIVs was evaluated using the F statistic, calculated as F = (beta/se)²^[16]^. An F value below 10 indicates a weak instrumental variable, potentially compromising the MR analysis. Therefore, only SNPs with adequate F values were retained.

If the initial threshold of P < 5 × 10⁻⁸ did not yield enough SNPs, it was relaxed to P < 5 × 10⁻⁶ ^[17]^ to ensure sufficient genetic variants for analysis, especially for less common exposures. Despite this adjustment, 7 skin diseases still lacked enough instrumental variables and were excluded from the analysis.

### 2.4. Statistical analysis

The study employed a range of advanced statistical methods, including Inverse Variance Weighted (IVW), MR-Egger regression, Weighted Median, Simple mode, and Weighted mode, to investigate the causal relationship between various skin diseases and non-small cell lung cancer (NSCLC). These methods provided a comprehensive analysis of potential genetic associations. The IVW method was used to estimate the overall causal effect by combining the effects of all selected SNPs, assuming no pleiotropy among genetic variants ^[18]^. MR-Egger regression, by relaxing the no-pleiotropy assumption, accounted for possible biases by allowing some instrumental variables to influence the outcome through pathways other than the exposure ^[19]^, The Weighted Median method, which requires that at least 50% of the total weight from genetic variants be valid ^[20]^. The Simple mode method offered stable estimates when dealing with a small number of instrumental variables, while the Weighted mode method reduced bias caused by heterogeneous effects among genetic variants ^[21]^. Cochran’s Q test assessed heterogeneity among SNPs, with a P < 0.05 indicating the presence of heterogeneity, suggesting that genetic variants may not be uniformly associated with the exposure or outcome. MR-Egger regression was also used to test for horizontal and directional pleiotropy, with SNPs causing significant pleiotropy (P < 0.05) being excluded to prevent bias ^[22]^. Finally, A leave-one-out analysis was conducted to systematically exclude each SNP individually, assessing the influence of each on the overall association between exposure and outcome, ensuring no single SNP disproportionately influenced the results.

## Results

### 3.1. Instrumental variable

In the dataset of 26 skin diseases selected for this study, 7 datasets still could not obtain sufficient instrumental variables even after lowering the inclusion and exclusion criteria. For 9 datasets, instrumental variables could only be obtained according to the reduced criterion of P < 5 × 10−6. The remaining 10 datasets were able to obtain instrumental variables according to the general standard. Among these, the number of instrumental variables for the skin diseases that yielded positive results were as follows:

Dermatitis herpetiformis: 6

Acne: 5

Hidradenitis suppurativa: 3

Rhinophyma: 4

Notably, Rhinophyma could only obtain instrumental variables after lowering the significance threshold to P < 5 × 10−6, whereas the SNPs used for the other positive results all met the criteria before the reduction. The positive results of the MR analysis are presented in Table 3, while the heterogeneity and horizontal pleiotropy tests are shown in Table 2.

**Table 1.** The information on skin diseases and non-small cell lung cancer data used in this study, all of which can be obtained from the FinnGen database, includes the following: Lung Cancers: Lung Squamous Cell Carcinoma (SCC), Lung Adenocarcinoma (ADC), Skin Diseases: Acne (AC), Hidradenitis Suppurativa (HS), Dermatitis Herpetiformis (DH), Infective Dermatitis (ID), Localized Scleroderma (LS), Cutaneous Abscess, Furuncle, and Carbuncle (CA), Atrophic Disorders of Skin (ADS), Papulosquamous Disorders (PSD), Atopic Dermatitis (AD), Urticaria (URT), Rhinophyma (RHN), Acquired Keratosis Palmaris et Plantaris (AKD), Pityriasis Lichenoides Chronica (PLC), Pityriasis Rosea (PR), Pyoderma (PYD), Xerosis Cutis (XER), Pruritus (PRU), Pemphigus (PEM), Seborrhoeic Dermatitis (SD), Pityriasis Rubra Pilaris (PRP), Lichen Planopilaris (LPP), Acne Conglobata (ACC), Erythema Multiforme (EM), Lupus Erythematosus (SLE), Chloasma (CHL), and Pigmented Purpuric Dermatosis (PPD).

| Trait | N.cases | N.controls | Populations | Year | Filtering threshold |
| --- | --- | --- | --- | --- | --- |
| SCC | 1787 | 345118 | European | 2024 | - |
| ADC | 2064 | 345118 | European | 2024 | - |
| AC | 3982 | 432686 | European | 2024 | $P < 5 \times 10^{-8}$ |
| HS | 1209 | 432686 | European | 2024 | $P < 5 \times 10^{-8}$ |
| DH | 585 | 451899 | European | 2024 | $P < 5 \times 10^{-8}$ |
| ID | 4195 | 394476 | European | 2024 | $P < 5 \times 10^{-8}$ |
| LS | 450 | 423041 | European | 2024 | $P < 5 \times 10^{-8}$ |
| CA | 10296 | 426987 | European | 2024 | $P < 5 \times 10^{-8}$ |
| ADS | 5037 | 423041 | European | 2024 | $P < 5 \times 10^{-8}$ |
| PSD | 16313 | 437420 | European | 2024 | $P < 5 \times 10^{-8}$ |
| AD | 26905 | 394476 | European | 2024 | $P < 5 \times 10^{-8}$ |
| URT | 12576 | 438050 | European | 2024 | $P < 5 \times 10^{-8}$ |
| RHN | 175 | 432686 | European | 2024 | $P < 5 \times 10^{-6}$ |
| AKD | 137 | 423041 | European | 2024 | $P < 5 \times 10^{-6}$ |
| PLC | 122 | 437420 | European | 2024 | $P < 5 \times 10^{-6}$ |
| PR | 242 | 437420 | European | 2024 | $P < 5 \times 10^{-6}$ |
| PYD | 643 | 426987 | European | 2024 | $P < 5 \times 10^{-6}$ |
| XER | 429 | 423041 | European | 2024 | $P < 5 \times 10^{-6}$ |
| PRU | 3819 | 394476 | European | 2024 | $P < 5 \times 10^{-6}$ |
| PEM | 208 | 451899 | European | 2024 | $P < 5 \times 10^{-6}$ |
| SD | 3382 | 394476 | European | 2024 | $P < 5 \times 10^{-6}$ |
| PRP | 71 | 437420 | European | 2024 | Insufficient data |
| LPP | 83 | 432686 | European | 2024 | Insufficient data |
| ACC | 175 | 432686 | European | 2024 | Insufficient data |
| EM | 754 | 438050 | European | 2024 | Insufficient data |
| SLE | 777 | 423041 | European | 2024 | Insufficient data |
| CHL | 127 | 423041 | European | 2024 | Insufficient data |
| PPD | 289 | 423041 | European | 2024 | Insufficient data |

**Table 2.** Pleiotropy and heterogeneity test of the MR positive results, including Dermatitis herpetiformis (DH), Acne (AC), Hidradenitis suppurativa (HS), and Rhinophyma (RHN). Lung Squamous Cell Carcinoma (SCC) and adenocarcinoma of the lung (ADC)

| Exposure | Outcome | Heterogeneity |  | Pleiotropy |
| --- | --- | --- | --- | --- |
|  |  | MR Egger Q (P) | IVW Q (P) | Egger intercept (P) |
| DH | SCC | 4.6051<br>(0.3302) | 4.9693<br>(0.4196) | 0.6038 |
| AC |  | 2.0474<br>(0.5626) | 2.9035<br>(0.5740) | 0.4230 |
| HS |  | 1.8801<br>(0.1703) | 10.1380<br>(0.0062) | 0.2834 |
| RHN |  | 0.4662<br>(0.7920) | 1.1637<br>(0.7617) | 0.4915 |
| DH | ADC | 4.5910<br>(0.3318) | 4.8008<br>(0.4406) | 0.6909 |
| AC |  | 0.9011<br>(0.8251) | 0.9226<br>(0.9212) | 0.8928 |
| HS |  | 0.0235<br>(0.8779) | 0.2673<br>(0.8748) | 0.7080 |
| RHN |  | 1.9837<br>(0.3708) | 2.1679<br>(0.5382) | 0.7095 |

**Table 3.**
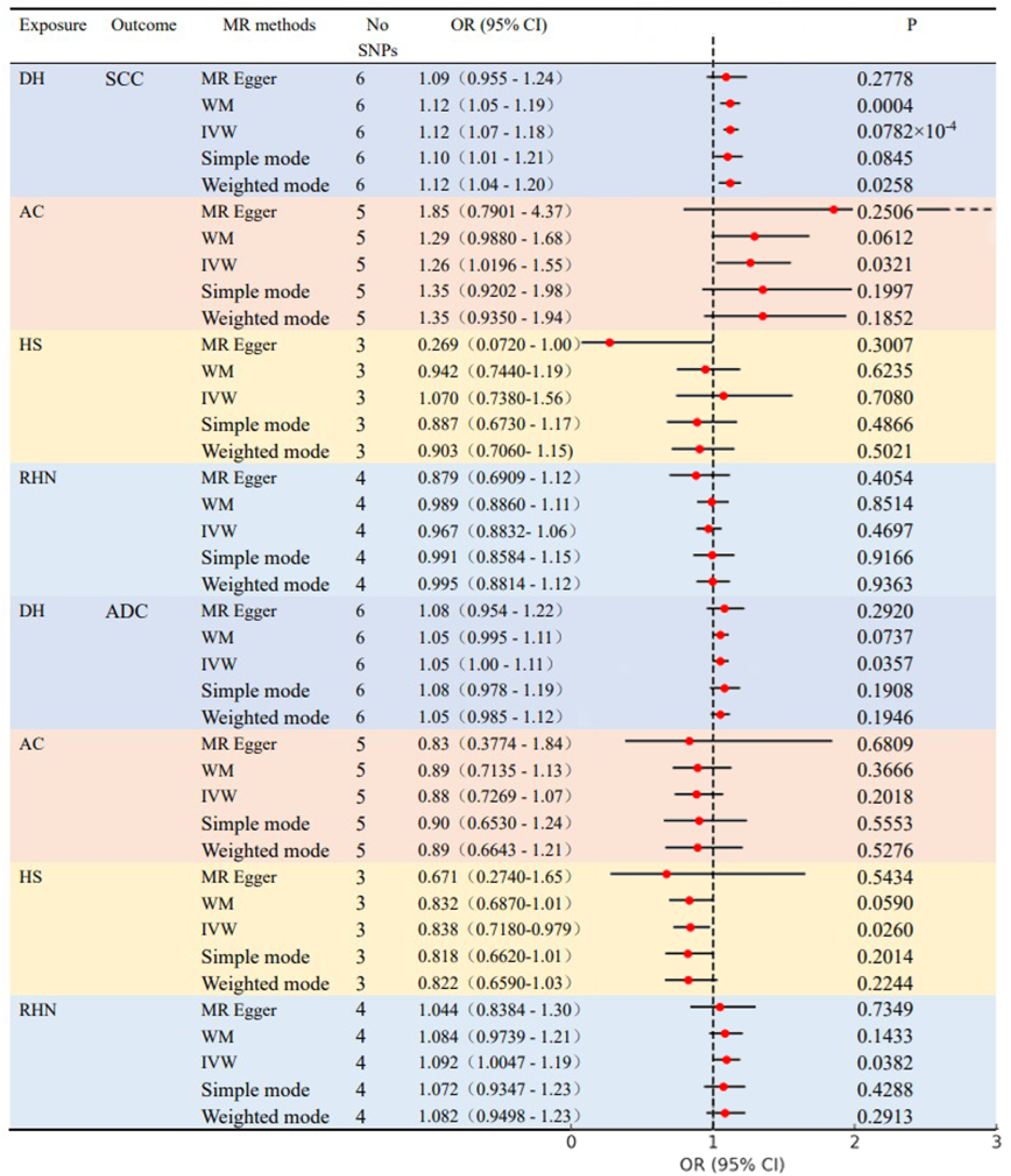
MR analysis results of skin diseases with positive associations and non-small cell lung cancer, including Dermatitis herpetiformis (DH), Acne (AC), Hidradenitis suppurativa (HS), and Rhinophyma (RHN), with Lung Squamous Cell Carcinoma (SCC) and adenocarcinoma of the lung (ADC).

#### 3.2.1. Dermatitis herpetiformis (DH) and NSCLC

Mendelian Randomization (MR) analyses revealed a significant causal relationship between dermatitis herpetiformis (DH) and squamous cell carcinoma (SCC) of the lung. The analysis demonstrated robust associations with p-values: IVW P = 0.0782 × 10⁻⁴, MR-Egger P = 0.2778, Weighted Median (WM) P = 0.0004, Simple mode P = 0.0845, and Weighted mode P = 0.0258. No heterogeneity was detected (Cochran’s Q test P > 0.05), and the MR-Egger intercept test confirmed no significant pleiotropy (P = 0.6038).

For adenocarcinoma (ADC), the causal association with DH was weaker, reflected in p-values: IVW P = 0.0357, MR-Egger P = 0.2920, WM P = 0.0737, Simple mode P = 0.1908, and Weighted mode P = 0.1946. Cochran’s Q test for ADC showed no heterogeneity (P > 0.05), and the MR-Egger intercept test confirmed no significant pleiotropy (P = 0.6909). Leave-one-out analysis confirmed that no single SNP disproportionately influenced the results. Scatter plots visually confirmed the consistency and direction of these findings across all methods. (Fig. 2).

**Figure 1.**
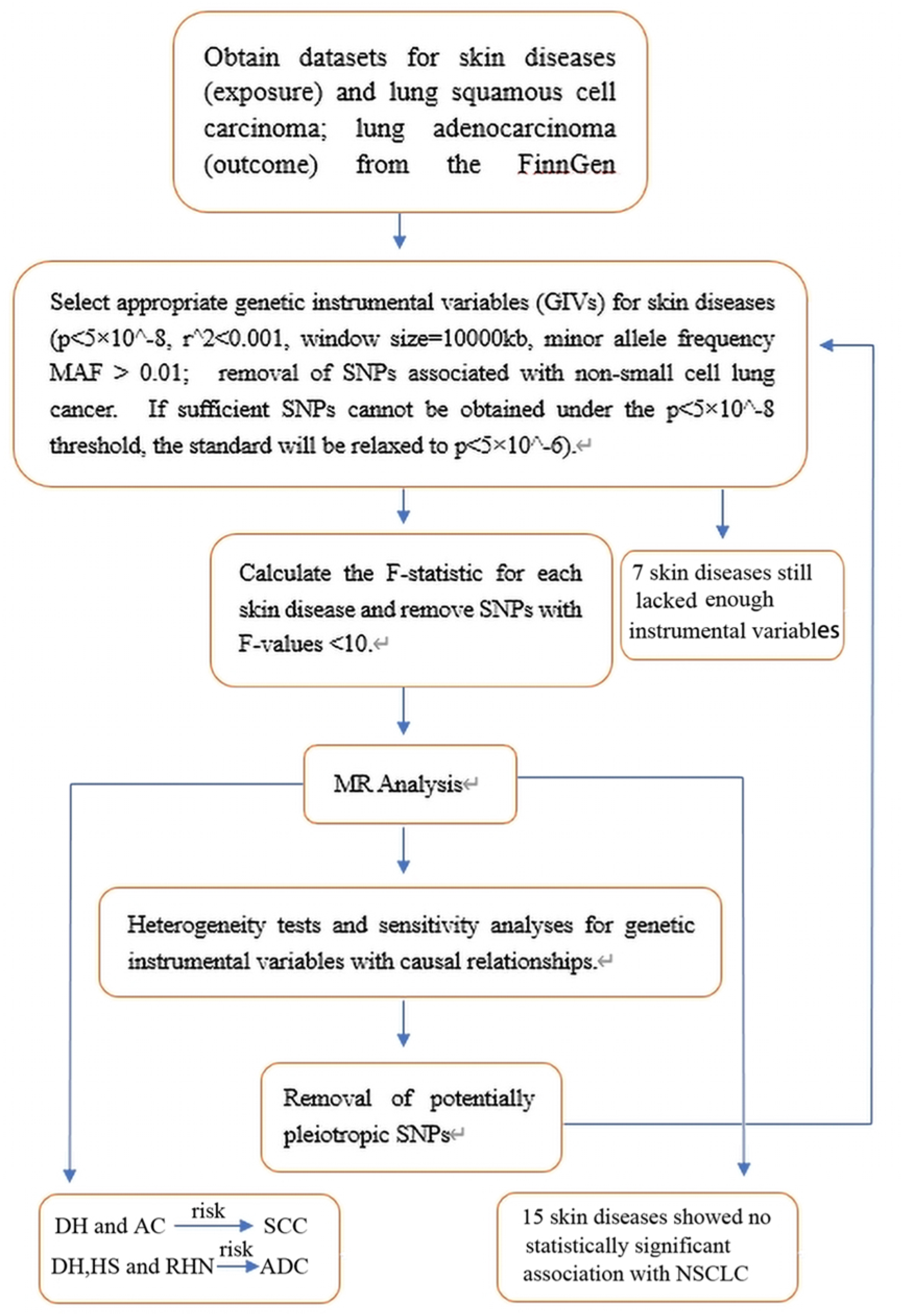
The flowchart of the study.

**Figure. 2.**
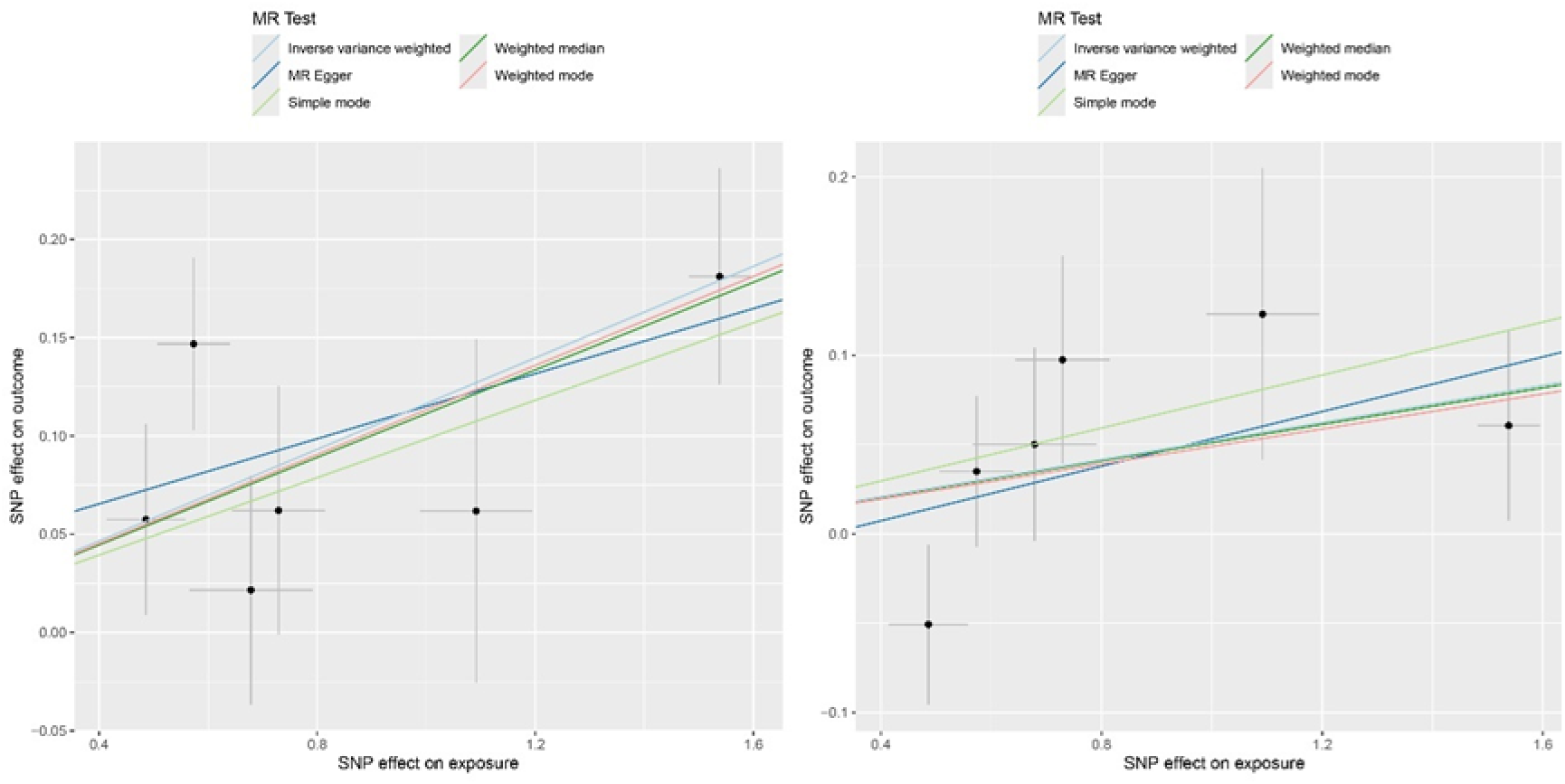
Dermatitis herpetiformis and NSCLC (SCC left ADC right)

#### 3.2.2. Acne (AC) and NSCLC

Mendelian Randomization (MR) analyses indicated a specific association between acne (AC) and squamous cell carcinoma (SCC) of the lung. P-values across methods were: IVW P = 0.0321, MR-Egger P = 0.2506, Weighted Median (WM) P = 0.0612, Simple mode P = 0.1997, and Weighted mode P = 0.1852. Cochran’s Q test (P > 0.05) confirmed no heterogeneity, and the MR-Egger intercept test (P = 0.4230) showed no significant pleiotropy, supporting the reliability of this association.

No causal relationship was found between AC and adenocarcinoma (ADC) of the lung, with all methods yielding p-values greater than 0.05. Cochran’s Q test (P > 0.05) and MR-Egger intercept test (P = 0.8928) confirmed no heterogeneity or pleiotropy, reinforcing the conclusion that AC is not related to ADC. Scatter plots visually depicted the causal relationship between AC and NSCLC (Fig. 3).

**Figure. 3.**
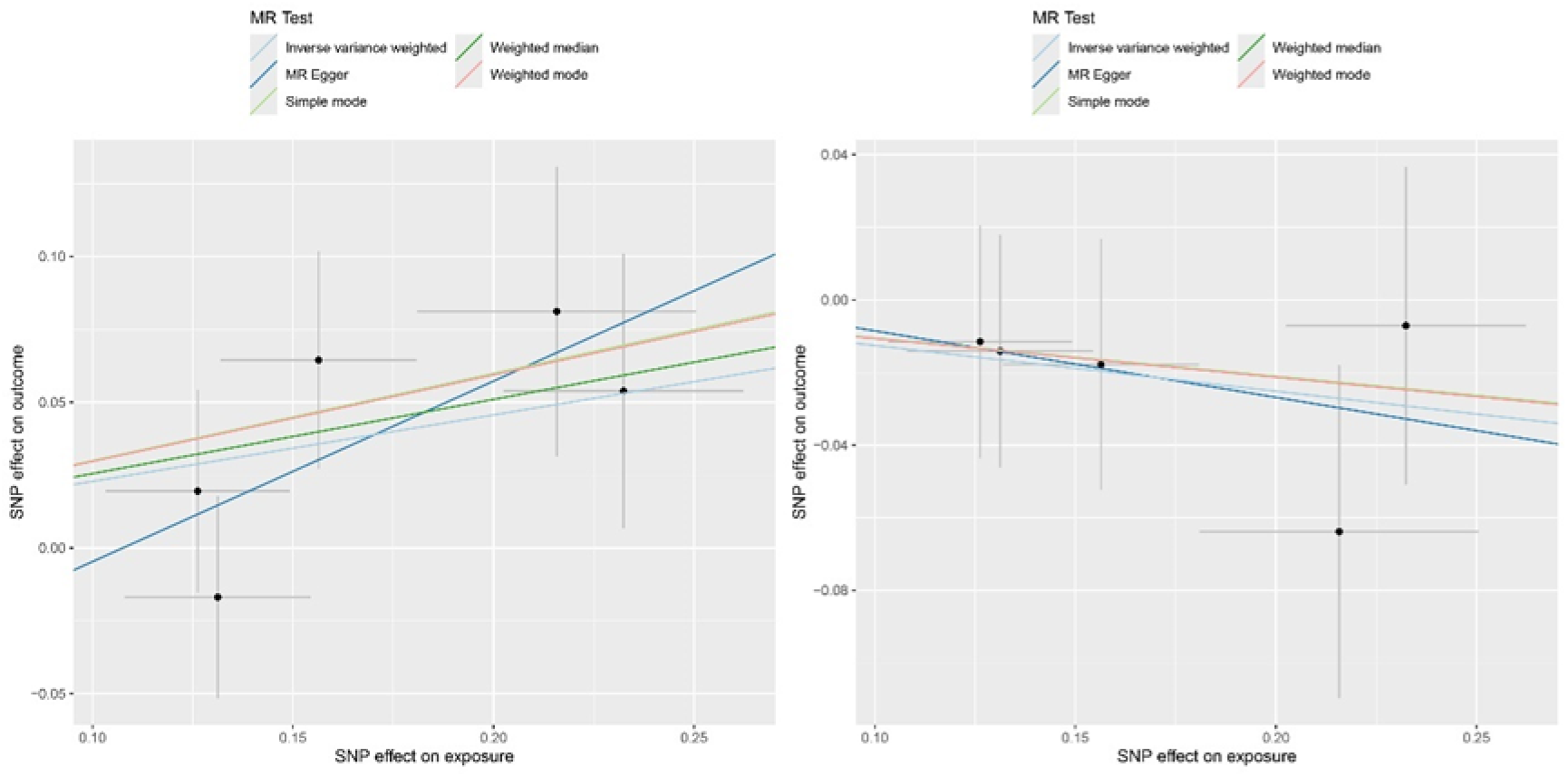
Acne and NSCLC (SCC left ADC right)

#### 3.2.3. Hidradenitis suppurativa (HS) and NSCLC

Mendelian Randomization (MR) analysis indicated a causal relationship between hidradenitis suppurativa (HS) and lung adenocarcinoma (ADC), supported by p-values: IVW P = 0.0260, MR-Egger P = 0.5434, Weighted Median (WM) P = 0.0590, Simple mode P = 0.2014, and Weighted mode P = 0.2244. Cochran’s Q test (P > 0.05) indicated no heterogeneity, and the MR-Egger intercept test (P = 0.7080) showed no significant pleiotropy, confirming the reliability of the HS- ADC association.

No causal relationship was found between HS and squamous cell carcinoma (SCC) of the lung, with all methods yielding p-values greater than 0.05. The MR-Egger intercept test (P = 0.2834) confirmed no significant pleiotropy, though the IVW Q test for SCC showed heterogeneity (P = 0.0062). This heterogeneity is less concerning due to the overall negative results for SCC. Leave-one-out analysis identified a single SNP with significant impact, but removing it left only two instrumental variables, insufficient for robust analysis. Scatter plots visually depicted the causal relationship between HS and NSCLC (Fig. 4).

**Figure. 4.**
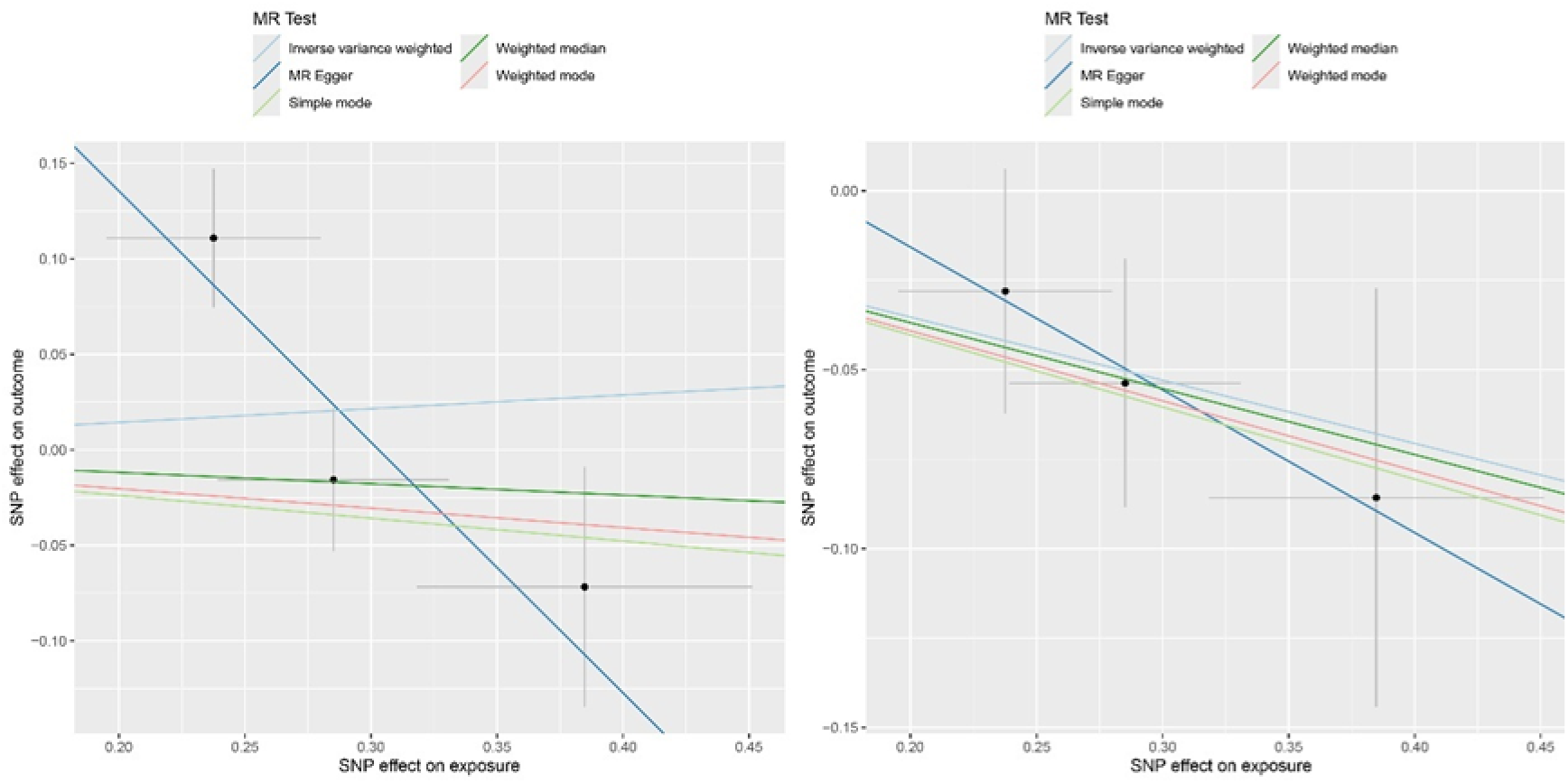
Hidradenitis suppurativa and NSCLC (SCC left ADC right)

#### 3.2.4. Rhinophyma (RHN) and NSCLC

Mendelian Randomization (MR) analyses suggested a potential causal relationship between rhinophyma (RHN) and adenocarcinoma (ADC) of the lung, with p-values: IVW P = 0.0382, MR-Egger P = 0.7349, Weighted Median (WM) P = 0.1433, Simple mode P = 0.4288, and Weighted mode P = 0.2913. Cochran’s Q test (P > 0.05) indicated no significant heterogeneity, and the MR-Egger intercept test (P = 0.7095) showed no significant pleiotropy.One SNP significantly impacted the results in the leave-one-out analysis, but only the IVW method retained a p-value below 0.05 after exclusion.

No causal relationship was found between RHN and squamous cell carcinoma (SCC), with all methods yielding p-values greater than 0.05. Cochran’s Q test (P > 0.05) and MR-Egger test (P = 0.4915) confirmed no heterogeneity or pleiotropy. Scatter plots illustrated these relationships (Fig. 5). The association between RHN and ADC should be interpreted with caution due to the reliance on a lowered significance threshold.

**Figure. 5.**
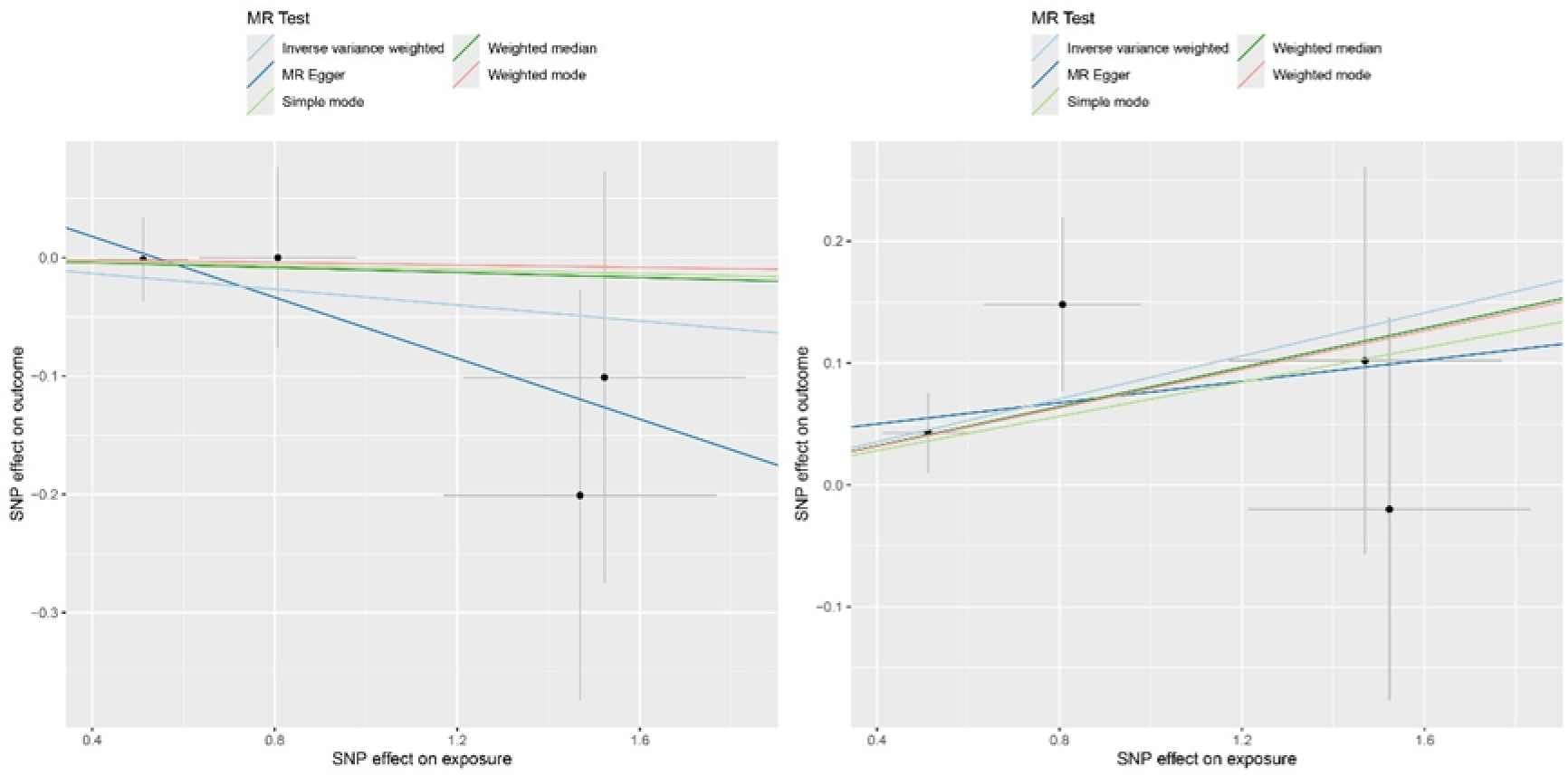
Rhinophyma and NSCLC (SCC left ADC right)

#### 3.2.5. The remaining 15 skin conditions and NSCLC

The remaining 15 skin conditions showed no association with the two types of NSCLC (all analysis methods P > 0.05). For datasets where the leave-one-out analysis revealed that certain SNPs interfered with the results, a new MR analysis was performed after removing the corresponding SNPs. The analysis conducted after removing the corresponding SNPs still failed to yield positive results.

## Discussion

In modern life sciences research, both the lungs and the skin originate from the mesoderm, indicating a shared homology in their growth and development ^[23]-[24]^. Pathophysiological studies have further confirmed the connection between the two. For example, in mucosal immunity, secretory immunoglobulin A (sIgA) serves as a key effector molecule. Research has shown its varying levels of expression in diseases such as asthma and chronic urticaria ^[25][26]^. Some studies focusing on the relationship between immunity, inflammation, and malignancy have identified key pathways, including Toll-like receptors (TLRs) and the tumor microenvironment (TME), which share inflammatory signals with skin diseases. For instance, cytokines such as TNF-α, IL-6, and IL-1β play critical roles in both lung and skin inflammation, suggesting that there may be mechanistic overlaps between pulmonary and dermatological conditions ^[6]^. In NSCLC, commonly used EGFR-TKI targeted drugs frequently cause skin rashes as the most common side effect. Current research on EGFR-TKI-related rashes suggests that the primary cause of these rashes is the disruption of the EGFR pathway, leading to interference with keratinocyte growth, migration, proliferation, and differentiation ^[27]^.

Recent MR studies on psoriasis and lung cancer published over the past two years ^[8]-[9]^ have bolstered our confidence in exploring this area further. Consequently, our team, building on these studies and leveraging the updated FinnGen R11 dataset from June 2024, selected 26 skin diseases as instrumental variables to investigate the relationship between skin diseases and non-small cell lung cancer.

Regarding the four skin diseases that yielded positive results: Dermatitis herpetiformis, Acne, Hidradenitis suppurativa, and Rhinophyma.

Dermatitis herpetiformis (DH) is characterized by blistering rashes, often accompanied by intense itching, and frequently co-occurs with celiac disease. Previous studies have identified a significant association between DH and the development of malignancies. A 2012 cohort study, with a 25-year follow-up, concluded that DH was significantly linked to the incidence of visceral and lymphatic tumors ^[28]-[29]^. However, few further in-depth studies have been conducted on this topic since then.

For Acne, there are a few reports linking it with malignancies such as breast cancer and prostate cancer. However, no other studies have clearly elucidated the relationship between Acne and lung cancer ^[30]-[31]^.

Hidradenitis suppurativa (HS) has been associated with lymphoma ^[32]^ and skin malignancies in previous studies. Approximately 0.5% to 4.6% of HS patients have later been diagnosed with cutaneous squamous cell carcinoma ^[33]^. Additionally, a systematic review encompassing 2,795 articles demonstrated an association between HS and psoriasis, indicating shared inflammatory pathways such as the IL-12-IL-23 axis, IL-17 interactions, and tumor necrosis factor (TNF)-α ^[34]^. Previous studies have confirmed the association between psoriasis and the risk of lung cancer ^[8]-[9]^. Therefore, the correlation between HS and NSCLC is supported by existing research.

Rhinophyma is generally considered an advanced manifestation of Rosacea. Currently, research specifically on Rhinophyma is limited, but studies on Rosacea have confirmed its association with an increased risk of skin and breast malignancies, particularly showing a significant correlation with HER2-negative breast cancer ^[35]-[36]^.

The MR analysis provides new evidence on the relationship between four skin diseases (Dermatitis herpetiformis, Acne, Rhinophyma, and Hidradenitis suppurativa) and the risk of NSCLC. However, the study has limitations, To ensure the consistency of the study population, we can currently only choose data with a predominantly European population. Additionally, although the causal relationships were analyzed using genetic instrumental variables, the results show that, apart from Dermatitis herpetiformis, the other three skin diseases are only associated with one type of NSCLC. Further research is needed to understand the specific relationships and patterns between different skin diseases and NSCLC subtypes. In addition, after the relevant genomic studies are further refined, further studies in Asian populations are needed to compare the results with the present study.

## Conclusion

This study confirms the association between four skin diseases and the risk of NSCLC. Dermatitis herpetiformis is significantly associated with an increased risk of squamous cell carcinoma of the lung. Acne shows a nominal association with an increased risk of squamous cell carcinoma of the lung. Rhinophyma, Hidradenitis suppurativa, and Dermatitis herpetiformis also exhibit nominal associations with an increased risk of adenocarcinoma of the lung. These findings provide direction for future clinical and experimental research related to NSCLC and highlight the importance of NSCLC screening and prevention in patients with these conditions. The fifteen skin diseases not found to be associated with NSCLC can help guide future research away from unrelated areas. Further genomic studies are necessary for the seven skin diseases for which sufficient SNP data were not obtained.

## Acknowledgments

We thank FinnGen database for their contributions.

## Data Availability

The dataset in this study can be found in the FinnGen database.

## Consent for publication

Not applicable.

